# Effect of Posterior Pericardiotomy for the Prevention of Postoperative Atrial Fibrillation in Off-pump Coronary Artery Bypass Grafting

**DOI:** 10.64898/2026.04.27.26351896

**Authors:** Suk Ho Sohn, Yoonjin Kang, Jae Woong Choi, Se Jin Oh, Ho Young Hwang

**Author notes:** **Corresponding Authors**, Ho Young Hwang, MD, PhD, Department of Thoracic and Cardiovascular Surgery, Seoul National University Hospital, Seoul National University College of Medicine, 101 Daehak-ro, Jongno-gu, Seoul 03080, Korea, (E-mail), Se Jin Oh, MD, PhD, Department of Thoracic and Cardiovascular Surgery, SMG-SNU Boramae Medical Center, Seoul National University College of Medicine, 20, Boramae-ro 5-gil, Dongjak-gu, Seoul 07061, Korea, (E-mail). Drs. Hwang HY and Oh SJ contributed equally to the study.

## Abstract

**Background:** This randomized controlled trial was conducted to evaluate the impact of posterior pericardiotomy on the prevention of postoperative atrial fibrillation (POAF) in patients who underwent off-pump coronary artery bypass grafting (CABG).

**Methods:** Adult patients who were scheduled to undergo isolated off-pump CABG were assessed for eligibility, and eligible patients were randomly assigned in a 1:1 manner to pericardiotomy or no intervention. Patients and postoperative caregivers were blinded to treatment assignment. The primary endpoint was the occurrence rate of POAF, and the secondary endpoints were the cumulative time spent in POAF and early outcomes, including operative mortality.

**Results:** A total of 403 patients were screened for eligibility, and 270 patients were randomly assigned to the posterior pericardiotomy group (n=136) or control group (n=134). The mean age was 67.8±10.3 years, and 20.4% (55 of 270 patients) were female. There was no intergroup difference in baseline characteristics or surgical data. Off-pump CABG was performed as planned in all patients except one. All patients received the assigned treatment regarding pericardiotomy, and no intergroup crossover occurred. POAF occurred in 27.8% (75 of 270 patients) of patients at a median of 2 days (interquartile range 1–3 days) after surgery. There was no significant difference in the occurrence rate of POAF between the two groups (30.1% and 25.4% in the pericardiotomy and control groups, *P* =.38). There were no significant differences in the secondary endpoints between the two groups.

**Conclusions:** Posterior pericardiotomy does not reduce the occurrence rate of POAF in patients undergoing off-pump CABG. (NCT06159985)

## INTRODUCTION

Postoperative atrial fibrillation (POAF) remains one of the most common complications following cardiac surgery, including coronary artery bypass grafting (CABG), with a reported rate of 10–53%.^1,2^ POAF usually occurs within the first week and peaks on the second postoperative day.^3^ POAF after CABG was previously considered benign, but recent evidence has shown that it is associated not only with early postoperative outcomes, including increased morbidity, prolonged hospital stay, and higher healthcare costs, but also with compromised long-term survival.^4^ To reduce the risk of POAF after CABG, several medical management practices, such as beta-blockers, have proven their efficacy and are currently incorporated into clinical practice.^5^ In addition, intraoperative posterior pericardiotomy (PP),^6^ which involves making an incision in the posterior pericardium and allowing drainage of any pericardial collection into the left pleural cavity, has been reported to decrease the occurrence rate of POAF after CABG.

Owing to the previous randomized controlled trials (RCTs) and meta-analyses and, most importantly, the PALACS (The Effect of Posterior Pericardiotomy on the Incidence of Atrial Fibrillation After Cardiac Surgery) trial,^7^ the ‘2023 ACC/AHA/ACCP/HRS Guideline for the Diagnosis and Management of Atrial Fibrillation’ recommend concomitant PP to be performed in patients undergoing CABG, aortic valve or ascending aortic aneurysm operations to reduce POAF incidence with a 2A class of recommendation.^8^ However, most of the existing evidence supporting the efficacy of PP has been derived from heterogeneous cohorts predominantly consisting of on-pump CABG or mixed cardiac surgical populations. The pathophysiology of POAF and the perioperative inflammatory milieu could differ substantially between on-pump CABG and off-pump CABG. In this context, an RCT entitled ‘Effect of Posterior Pericardiotomy for the Prevention of Postoperative Atrial Fibrillation in Off-pump Coronary Artery Bypass Grafting (ELIMINATE-AF)’ was conducted to evaluate the impact of PP on the prevention of POAF exclusively in patients undergoing off-pump CABG (ClinicalTrials.gov identifier: NCT06159985).

## MATERIALS AND METHODS

### Study Design

The ELIMINATE-AF trial was designed as an RCT involving 3 surgeons from 2 institutions. The institutional review board in each hospital approved the study protocol (approval date 10/25/2023; approval no. H-2309-096-1469 and 30-2023-101), and informed consent was obtained from all the study participants. The study was designed in accordance with the Consolidated Standards of Reporting Trials statement.^9^ Patients over 19 years of age who were scheduled to undergo primary isolated off-pump CABG for multivessel disease on a nonemergency basis were assessed for eligibility for study enrollment. The exclusion criteria included patients with (1) planned on-pump CABG; (2) concomitant cardiac procedures, including valve or aorta surgery; (3) a history of paroxysmal, persistent, or permanent atrial fibrillation (AF); (4) no use of the left internal thoracic artery (LITA) as a graft; (5) left pleural disease prohibiting left pleural opening; (6) minimally invasive direct coronary artery bypass (MIDCAB); (7) critical preoperative status; (8) previous cardiac surgery; (9) immunosuppression; (10) pericardial disease; and (11) those who refused to participate in the study.

Between December 2023 and December 2025, 403 patients were assessed for eligibility, and 127 patients were excluded. Of the 276 patients eligible for the study, 6 refused to participate, and 270 were enrolled. The enrolled patients were randomly assigned to the posterior pericardiotomy group (the PP group) or the no intervention group (the Control group) in a 1:1 ratio (Figure 1).

**FIGURE 1.**
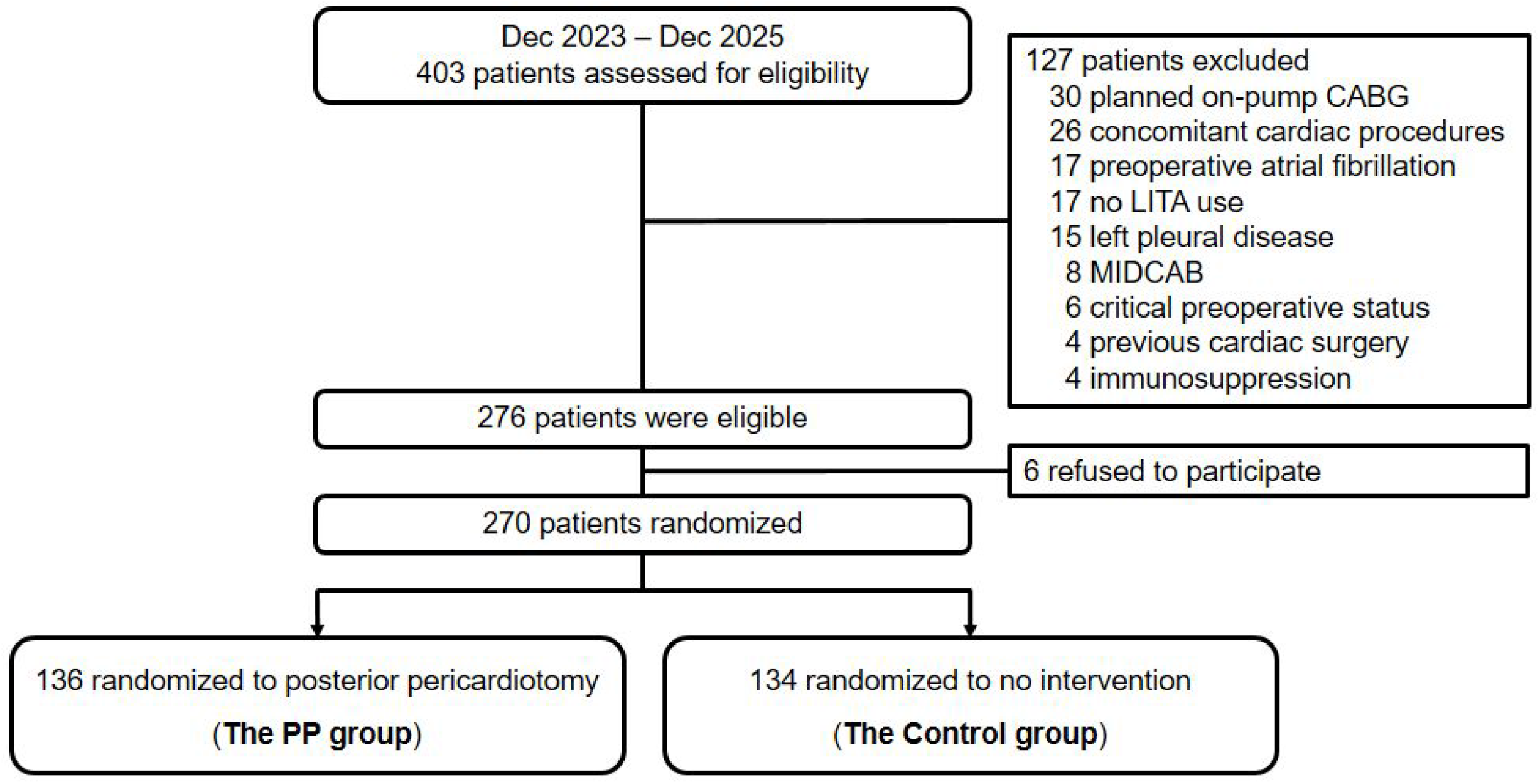
Flow diagram of the study population. CABG, coronary artery bypass grafting; LITA, left internal thoracic artery; MIDCAB, minimally invasive direct coronary artery bypass.

### Operative Strategies and Randomization Process

Off-pump CABG was planned for all isolated CABG patients regardless of left ventricular function during the study period. The in situ LITA was harvested using a skeletonization or semi-skeletonization technique. The SV was harvested simultaneously using a no-touch technique that avoided luminal dilatation and retained perivascular soft tissue, as previously described.^10^ Randomization was performed after the absence of left pleural adhesion was confirmed during the LITA harvest and the creation of posterior pericardiotomy was feasible. Web-based block randomization was performed using randomly determined block sizes of 4 and 6. After randomization, the CABG procedures were performed as planned. For multivessel revascularization, Y-composite grafting based on in situ LITA or aortocoronary SV sequential grafting in addition to LITA-LAD anastomosis was preferred. After completion of the revascularization procedure, the patients assigned to the PP group received a 5–6 cm vertical incision posterior to the phrenic nerve and extending from the left inferior pulmonary vein to the diaphragm.^7^ No intervention was performed in the Control group.

### Postoperative management

Patients and postoperative caregivers were blinded to treatment group assignment. Patients in both groups were prescribed routine pre- and postoperative beta-blockers. During the hospital stay, continuous electrocardiogram (ECG) monitoring was performed for all patients until discharge, and 12-lead ECGs were also assessed on a daily basis. If any abnormal findings were detected or suspected during continuous ECG monitoring, a 12-lead ECG was acquired and documented to confirm the occurrence of any arrhythmia. If POAF was detected, an anti-arrhythmic drug (e.g., amiodarone) was used. Systemic anticoagulation was initiated if POAF persisted for more than 24 hours.

### Evaluation of Endpoints

The primary endpoint of the ELIMINATE-AF trial was the occurrence rate of POAF, which was defined as the documentation of an irregular atrial rhythm without discrete P waves that lasted ≥30 seconds during the postoperative period.^8^ The secondary endpoints were the time to onset of POAF, which was defined as the time from the surgery end to the first documentation of POAF; the cumulative time spent in POAF, which was defined as the sum of the durations of POAF during the postoperative hospital stay measured by continuous ECG monitoring; and early postoperative outcomes, including operative mortality, delirium, respiratory complications, acute kidney injury, bleeding reoperation, stroke, mediastinitis, and length of hospital stay. The incidence of cardiac tamponade was also assessed.

### Statistical Analysis

The null hypothesis was that the occurrence rate of POAF would be lower in the PP group than in the Control group. Based on previous studies, the occurrence rate of POAF was estimated to be 30.0% in the Control group and 15.0% in the PP group.^7^ With a superiority design, this study was designed to have 80% power to detect a significant difference in the occurrence rate of POAF between the two groups, with a two-sided type I error of 5.0%. According to the power calculation, 121 patients were needed in each group. To account for a 10% dropout rate, we planned to recruit a total of 270 patients, with 135 patients in each group.

Statistical analyses were performed using SPSS software (version 29.0; IBM, Armonk, NY, USA) and SAS software (version 9.3; SAS Institute, Cary, NC, USA). Continuous variables are presented as the means ± standard deviations for normally distributed variables and as medians with interquartile range (IQR) for non-normally distributed variables. Categorical variables are presented as counts and percentages of subjects. Comparisons between the two groups were performed using the chi-square test or Fisher’s exact test for categorical variables and Student’s t test or the Wilcoxon rank sum test for continuous variables. Sensitivity analysis for the primary endpoint was performed using a generalized linear mixed model that included the operating surgeon as a random effect and the following covariates as fixed effects: age, sex, smoking status, body mass index >25.0 kg/m^2^, hypertension, diabetes mellitus, dyslipidemia, history of stroke, dialysis, chronic obstructive pulmonary disease, peripheral arterial occlusive disease, previous percutaneous coronary intervention, left ventricular ejection fraction ≤ 0.35, preoperative diagnosis, three-vessel disease, and left main disease. Subgroup analyses were performed to investigate the following possible effect modifiers: age (<70 years vs. ≥70 years), sex, left ventricular ejection fraction ≤ 0.35, and preoperative diagnosis. Secondary endpoints were analyzed using descriptive statistics and relative risks.

A *P* value of <0.05 was considered to indicate statistical significance. All outcomes were analyzed in an intention-to-treat base.

## RESULTS

### Baseline Characteristics

The mean age of the patients was 67.8 ± 10.3 years, and 20.4% of the patients (55 out of 270 patients) were female. The demographic data and preoperative risk factors were not significantly different between the two groups (Table 1).

**TABLE 1.**
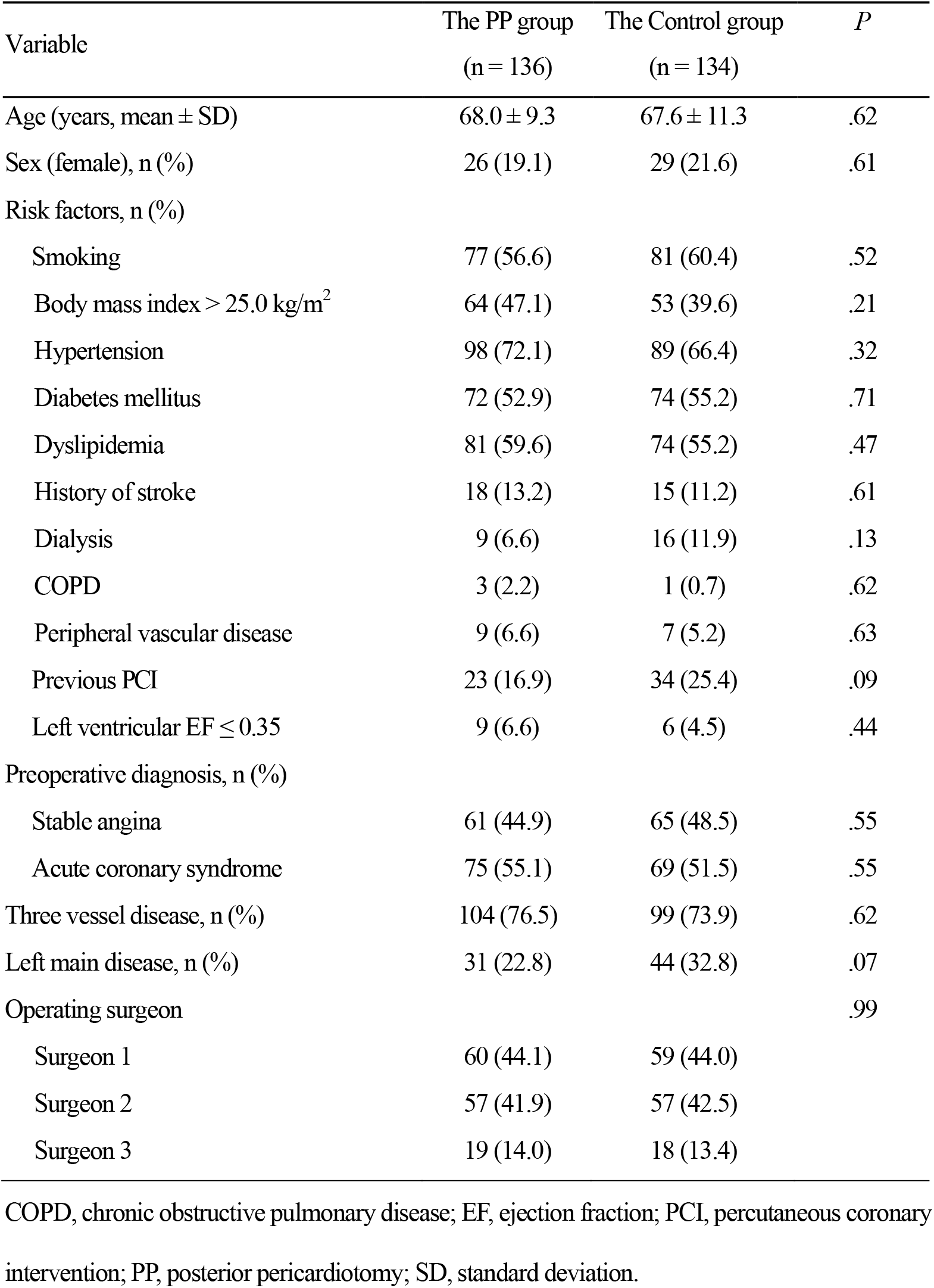
Preoperative characteristics and risk factors for the study patients.

| Variable | The PP group<br>(n = 136) | The Control group<br>(n = 134) | *P* |
| --- | --- | --- | --- |
| Age (years, mean ± SD) | 68.0 ± 9.3 | 67.6 ± 11.3 | .62 |
| Sex (female), n (%) | 26 (19.1) | 29 (21.6) | .61 |
| Risk factors, n (%) |  |  |  |
| Smoking | 77 (56.6) | 81 (60.4) | .52 |
| Body mass index $> 25.0$ kg/m<sup>2</sup> | 64 (47.1) | 53 (39.6) | .21 |
| Hypertension | 98 (72.1) | 89 (66.4) | .32 |
| Diabetes mellitus | 72 (52.9) | 74 (55.2) | .71 |
| Dyslipidemia | 81 (59.6) | 74 (55.2) | .47 |
| History of stroke | 18 (13.2) | 15 (11.2) | .61 |
| Dialysis | 9 (6.6) | 16 (11.9) | .13 |
| COPD | 3 (2.2) | 1 (0.7) | .62 |
| Peripheral vascular disease | 9 (6.6) | 7 (5.2) | .63 |
| Previous PCI | 23 (16.9) | 34 (25.4) | .09 |
| Left ventricular EF $\leq 0.35$ | 9 (6.6) | 6 (4.5) | .44 |
| Preoperative diagnosis, n (%) |  |  |  |
| Stable angina | 61 (44.9) | 65 (48.5) | .55 |
| Acute coronary syndrome | 75 (55.1) | 69 (51.5) | .55 |
| Three vessel disease, n (%) | 104 (76.5) | 99 (73.9) | .62 |
| Left main disease, n (%) | 31 (22.8) | 44 (32.8) | .07 |
| Operating surgeon |  |  | .99 |
| Surgeon 1 | 60 (44.1) | 59 (44.0) |  |
| Surgeon 2 | 57 (41.9) | 57 (42.5) |  |
| Surgeon 3 | 19 (14.0) | 18 (13.4) |  |

COPD, chronic obstructive pulmonary disease; EF, ejection fraction; PCI, percutaneous coronary intervention; PP, posterior pericardiotomy; SD, standard deviation.

### Operative Data

All patients received the assigned treatment, and no intergroup crossover occurred. All but one patient underwent off-pump CABG as planned. On-pump conversion was performed in the other patient due to hemodynamic instability during obtuse marginal branch anastomosis. The average number of distal anastomoses per patient was 3.7 ± 1.0, and there were no intergroup differences (*P* =.92). The average number of distal anastomoses per LITA, per RITA, and per SV was 1.0 ± 0.2, 0.1 ± 0.5, and 2.6 ± 1.1, respectively, without any intergroup differences (Table S1).

### Primary and Secondary Endpoints

POAF occurred in 27.8% (75 of 270 patients) of the patients at a median of 2 (IQR 1, 3) days after surgery. There was no significant difference in the occurrence rate of POAF between the two groups (30.1% [41 of 136 patients] and 25.4% [34 of 134 patients] in the PP and Control groups, respectively; *P* =.38). The sensitivity analysis for the primary endpoint using a generalized linear mixed model also revealed no significant difference in the occurrence rate of POAF between the two groups (adjusted relative risk [95% confidence interval] = 1.27 (0.78, 2.06), *P* =.34). The treatment effect was similar across all prespecified subgroup analyses (Figure 2).

**FIGURE 2.**
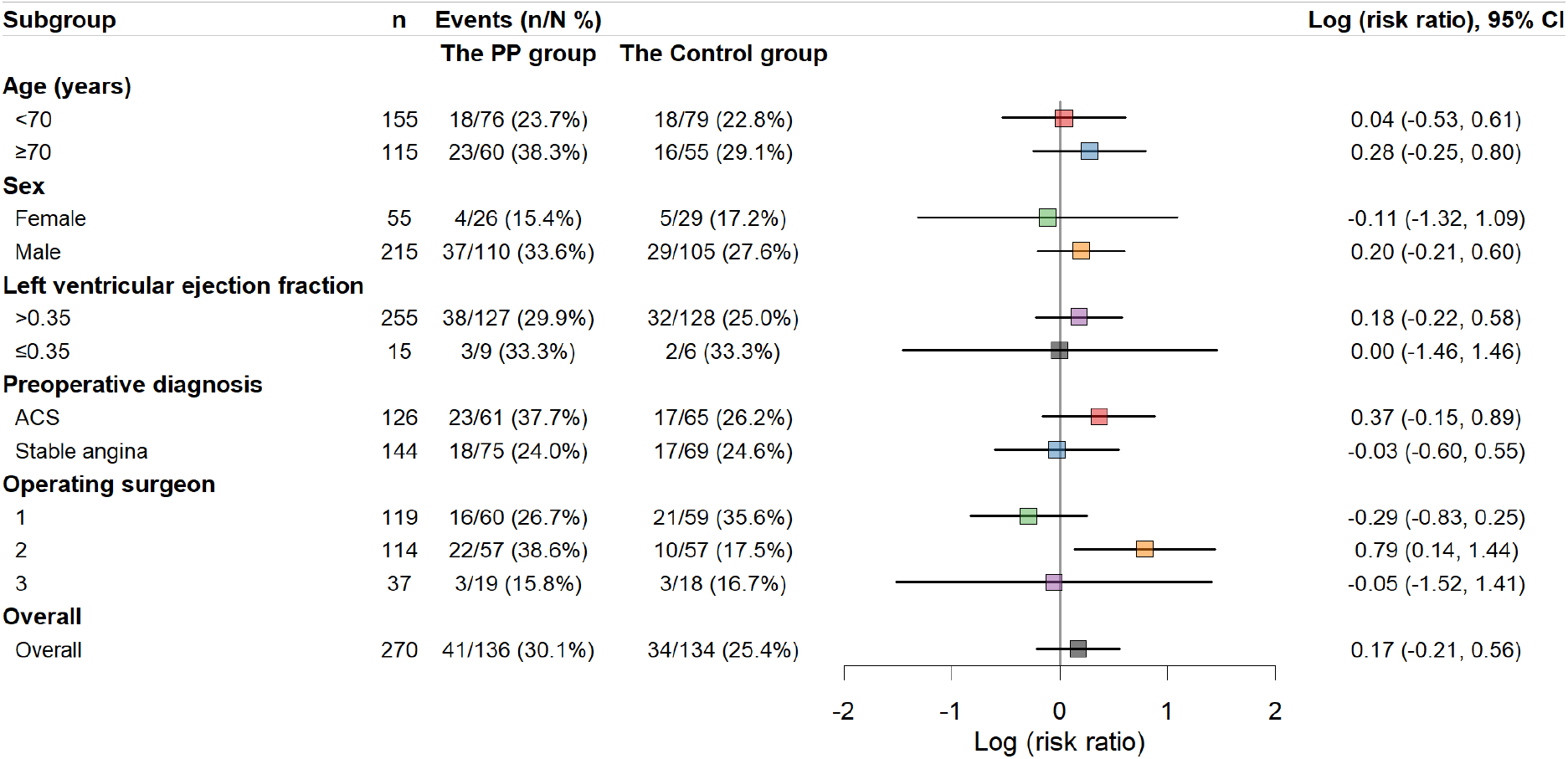
Subgroup analysis for the primary endpoint. The treatment effect was similar across all the prespecified subgroup analyses except for the effect of the operating surgeon.

The median time to POAF onset was 37.3 (21.8, 76.6) hours and 49.4 (32.7, 64.7) hours in the PP and Control groups, respectively (*P* =.13) (Figure 3). The cumulative times in the AF were 9 (4, 14) hours and 5 (1, 24) hours in the PP and Control groups, respectively (*P* = .22). There were no significant differences in other secondary endpoints, including the median length of postoperative hospital stay (7 days [IQR 7, 9]), operative mortality or postoperative complications, between the two groups (Table 2).

**TABLE 2.** Primary and secondary endpoints.

| Variables | Total<br>(n = 270) | The PP group<br>(n = 136) | The Control group<br>(n = 134) | *P* |
| --- | --- | --- | --- | --- |
| Primary endpoint |  |  |  |  |
| Postoperative AF | 75 (27.8) | 41 (30.1) | 34 (25.4) | .38 |
| Secondary endpoints |  |  |  |  |
| Operative mortality | 1 (0.4) | 1 (0.7) | 0 (0.0) | >.99 |
| Delirium | 17 (6.3) | 10 (7.4) | 7 (5.2) | .47 |
| Respiratory complications | 9 (3.3) | 5 (3.7) | 4 (3.0) | >.99 |
| Acute kidney injury | 4 (1.5) | 2 (1.5) | 2 (1.5) | >.99 |
| Bleeding reoperation | 3 (1.1) | 1 (0.7) | 2 (1.5) | .62 |
| Stroke | 2 (0.7) | 0 (0.0) | 2 (1.5) | .25 |
| Mediastinitis | 0 (0.0) | 0 (0.0) | 0 (0.0) | - |
| Cardiac tamponade | 0 (0.0) | 0 (0.0) | 0 (0.0) | - |
| Cumulative time in AF (hours) | 7 (2, 15) | 9 (4, 14) | 5 (1, 24) | .22 |
| Hospital stay (days) | 7 (7, 9) | 7 (7, 9) | 7 (7, 9) | .47 |

Values are presented as n (%) or medians with interquartile ranges.
AF, atrial fibrillation; PP, posterior pericardiotomy.

**FIGURE 3.**
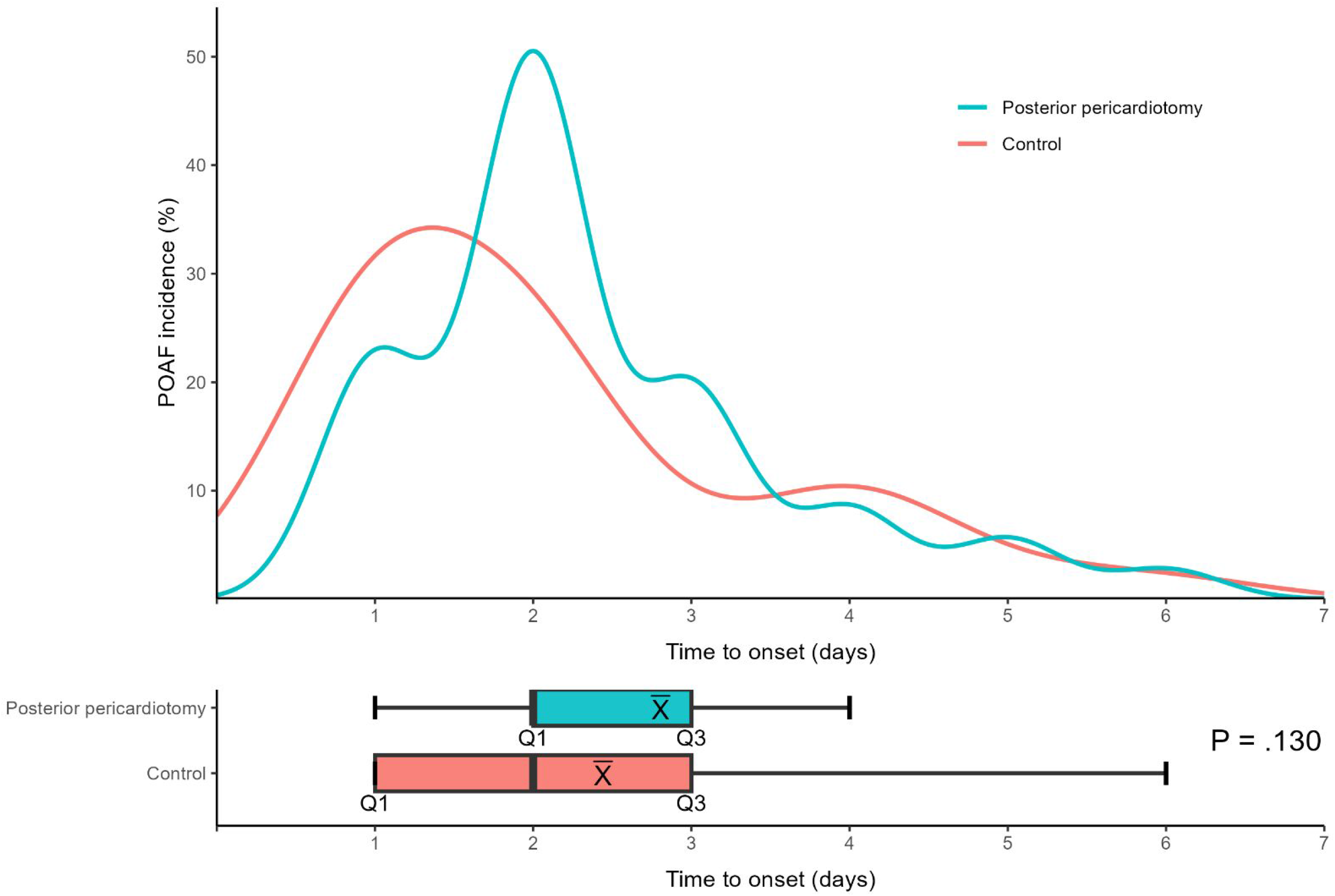
Comparison of the time to onset of POAF between the posterior pericardiotomy group and the control group. In the box plot, the dashed line represents the median, and X represents the mean. Q1, lower quartile; Q3, upper quartile.

## DISCUSSION

The present study demonstrated that PP did not significantly reduce POAF after off-pump CABG. These findings suggest that the preventive effect of PP observed in on-pump cardiac surgery may not be readily extrapolated to the off-pump CABG population.

The pathogenesis of POAF following cardiac surgery is multifactorial. One mechanism is the initiation of arrhythmia triggered by a susceptible substrate, including structural remodeling, atrial fibrosis, apoptosis, increased oxidative stress, and metabolic changes after CABG.^11^ Another mechanism is that pericardial effusion, a known post-CABG complication that is prominently collected in the left posterior space, can establish AF by causing mechanical irritation, inflammation, and increased atrial pressure, which induces atrial stretching and disrupts electrical conduction, promoting ectopic activity and re-entry circuits.^12,13^ An increasing amount of evidence supports the concept that shed mediastinal blood within the pericardium might exert not only the direct mechanical compression but also foster a highly pro-oxidant and pro-inflammatory milieu rich in leucocytes in proximity to the atria that could in turn trigger POAF through breakdown products, activation of the coagulation cascade and oxidative bursts.^13-15^ Clinical and experimental studies have suggested that POAF can be triggered even by small amounts of pericardial effusion, causing localized inflammation.^13^

PP was first described in 1995 as a simple procedure to drain fluid from the pericardial space into the left pleural space and was demonstrated to reduce postoperative pericardial effusion and related arrhythmias in CABG patients.^6^ The effects of PP on the reduction of pericardial effusion and its association with POAF have been evaluated in previous RCTs, and several meta-analyses that incorporated these RCTs have also been published. A previous study^16^ pooled 18 RCTs (3531 patients) and found that PP was associated with a significantly lower occurrence rate of POAF (OR 0.45, 95% CI 0.32–0.64; *P* < 0.0001), as well as a lower incidence of early and late pericardial effusion and cardiac tamponade. Another meta-analysis^17^ included 25 RCTs comparing PP versus no intervention (4467 patients) and revealed that the overall occurrence rate of POAF was 11.7% in the PP group compared with 23.7% in the control group, with a significant decrease in the risk of POAF in the intervention group (OR 0.49, 95% CI 0.38–0.61). However, when pooling the related studies by geographical regions, the benefits of PP remained evident only in RCTs performed in Turkey and Egypt, whereas those conducted in North America, Europe, and Asia failed to reach statistical significance, although a trend favoring PP was present.^18^

To overcome the limitations of underpowered and/or poorly designed RCTs, the PALACS (The Effect of Posterior Pericardiotomy on the Incidence of Atrial Fibrillation After Cardiac Surgery) trial^7^ was conducted with adequate power to compare PP versus no intervention. It found that the incidence of POAF was significantly lower in the PP group compared to the non-PP group (17% vs. 33%), with a relative risk of 0.55. Although this study included more than 400 patients, they enrolled a mixed patient population including patients who underwent CABG, aortic valve, and aortic procedures, and the reduction of POAF in the CABG-only subgroup did not reach statistical significance (19% vs. 25%). Furthermore, there might be differences in the mechanism of POAF in the off-pump CABG population in which the detrimental effects of cardiopulmonary bypass, hypothermia and cardioplegic arrest of the heart could be avoided. Only one previous RCT specifically evaluated the possible benefit of PP in off-pump CABG patients.^19^ In this study, which randomized 207 patients to the PP group (n = 105) and the control group (n = 102), the authors demonstrated that the presence of PP had no effect on the rate of occurrence of POAF among off-pump CABG patients (4.8% vs. 5.9% in the PP group vs. control group, *P* =.719). However, this study is limited in that continuous monitoring was applied only during the first 48 hours, and that an arrhythmia sustained for more than 30 minutes was considered significant.

The present study has several strengths: (1) 24-hour tele-monitoring was applied until discharge in all patients; (2) a homogenous population received off-pump CABG, except one; and (3) pre- and postoperative beta-blockers were routinely prescribed. The results of this trial demonstrated that the benefit of PP may be limited in patients undergoing isolated off-pump CABG. This limited benefit of PP in off-pump CABG might be due to the lower arrhythmogenic triggers and less mediastinal shed blood caused by avoiding the use of cardiopulmonary bypass. In addition, the routine use of pre- and postoperative beta-blockers might have attenuated the effect of PP in this study. Although the occurrence rate of POAF in the present study was relatively higher (27.8% of the entire population) than that reported in previous studies, this could be largely attributed to the fact that the mean age of the enrolled patients, approximately 68 years, was older than that in previous studies, which ranged from 61 to 64.^1,7^ In addition, a simple comparison of the occurrence rate of POAF to other studies is not appropriate, as patients characteristics, particularly those related to regional and ethnic variability, may differ between studies. Finally, meticulous ECG monitoring, including 24-hour tele-monitoring conducted from the operating room until just before discharge to capture every POAF in every patient and 12-lead ECG on a daily basis, would affect the high occurrence rate of POAF in the present study. The authors consider the occurrence rate of POAF of 27.8% in this trial to be reliable, as previous retrospective studies conducted at our institution reported rates of 33.7-34.7%.^20,21^

### Limitations

The present study has several limitations. First, although this study was designed as a randomized controlled trial and power calculations were strictly performed, the sample size was relatively small. Second, the amount of postoperative pericardial effusion was not quantitatively assessed, limiting mechanistic interpretation.

## Conclusion

Posterior pericardiotomy does not reduce the occurrence rate of POAF in patients undergoing off-pump CABG.

## Data Availability

Upon request, deidentified participant data could be shared at the discretion of the corresponding author (HYH) by e-mail contact.

## Abbreviations

AF: atrial fibrillation
CABG: coronary artery bypass grafting
ECG: electrocardiogram
IQR: interquartile range
LAD: left anterior descending artery
LCX: left circumflex artery
LITA: left internal thoracic artery
MIDCAB: minimally invasive direct coronary artery bypass
PALACS: The Effect of Posterior Pericardiotomy on the Incidence of Atrial Fibrillation After Cardiac Surgery
POAF: postoperative atrial fibrillation
PP: posterior pericardiotomy
RCA: right coronary artery
RCT: randomized controlled trial
RITA: right internal thoracic artery
SV: saphenous vein

## Acknowledgments

The authors thank the Medical Research Collaborating Center, Seoul National University Hospital, for the statistical analyses and consultation.

